# A molecular biofluid signature of multiple system atrophy (MSA): CSF neurofilament light chain and α-synuclein seeding as complementary biomarkers allow to distinguish MSA from sporadic adult-onset ataxia

**DOI:** 10.64898/2026.05.11.26352905

**Authors:** Vaibhavi Kadam, Luis Concha-Marambio, Lukas Beichert, Anja Heider, Thomas Klockgether, Jennifer Faber, Kathrin Brockmann, Ludger Schoels, Benjamin Roeben, David Mengel, Matthis Synofzik

**Affiliations:** Division Translational Genomics of Neurodegenerative Diseases, Hertie Institute for Clinical Brain Research and Center of Neurology, University of Tuebingen, Tuebingen, Germany; German Center for Neurodegenerative Diseases (DZNE) Tuebingen, Tuebingen, Germany; Graduate School of Cellular and Molecular Neuroscience, University of Tuebingen, Tuebingen, Germany; Amprion, R&D Department, San Diego, California, USA; German Center for Neurodegenerative Diseases (DZNE) Bonn, Bonn, Germany; Department of Parkinson, Sleep and Movement Disorders, Center for Neurology, University Hospital Bonn, Bonn, Germany; Department for Neuroradiology, University Hospital Bonn, Bonn, Germany; Department of Neurodegenerative Diseases, Hertie Institute for Clinical Brain Research and Center of Neurology, University of Tuebingen, Tuebingen, Germany

**Keywords:** multiple system atrophy, sporadic adult-onset ataxia, α-synuclein, seed amplification assay, neurofilament light chain, cerebrospinal fluid, biological diagnosis, diagnosis

## Abstract

**Background:** Accurate diagnosis of multiple system atrophy (MSA) is critical for clinical management and efficient trial designs, yet remains challenging, particularly distinguishing MSA (especially cerebellar-subtype [MSA-C]) from sporadic adult-onset ataxia (SAOA). Combining a marker of neuroaxonal degeneration, neurofilament light chain (NfL), with a marker of the pathogenic MSA hallmark, α-synuclein seeding activity, may define a mechanistically-informed CSF signature of MSA, enabling sensitive and specific differentiation from SAOA even in early disease.

**Methods:** We analyzed 60 cross-sectional patient CSF samples (n=32 clinically diagnosed MSA [MSA_clin_] 22/32 MSA-C; n=28 SAOA) for NfL (Simoa) and α-synuclein seeding activity (seed amplification assay [synSAA], Piperazine-N,N’-bis(2-ethanesulfonic acid)-based), and assessed diagnostic accuracy, disease-duration correlations, and trial power using biomarker-based stratification.

**Results:** Age-adjusted NfL was higher in MSA_clin_ than SAOA (3859 vs. 997pg/mL), yielding 96.9% sensitivity and 85.7% specificity. SynSAA was concordant with clinical diagnosis (25/32 MSA_clin_ synSAA-positive; 23/28 SAOA synSAA-negative), with 78.1% sensitivity and 85.2% specificity (all confirmed in MSA-C subgroup). Both biomarkers displayed divergent trajectories with disease duration: NfL peaked early before declining (r=–0.45, p=0.01); whereas synSAA maximum fluorescence intensity increased (r=0.42, p=0.016), suggesting greater synSAA signal with accumulating MSA burden. Integrating both biomarkers in MSA treatment trials allows sample-size reduction by 20% versus NfL alone.

**Conclusions:** CSF NfL and synSAA capture complementary aspects of MSA biology: while NfL provides high diagnostic accuracy for MSA_clin_, peaking early, synSAA adds mechanistic specificity for α-synuclein seeding activity and might allow target engagement assessment. Combined, they might enable biological diagnostic frameworks, molecular trial stratification, and treatment monitoring in MSA.

**Key messages:** *What is already known on this topic:* While highly warranted for clinical management and efficient treatment trial design, accurate diagnosis of multiple system atrophy (MSA) against overlapping and reciprocally mimicking conditions such as sporadic adult-onset ataxia (SAOA) remains clinically challenging, especially in early disease stages. A mechanistically informed biofluid signature of MSA might enable sensitive and specific differentiation from SAOA, even in early disease stage. Recently merging molecular markers reflecting neuroaxonal damage (NfL) and α-synuclein seeding activity (measured by the seed amplification assay; synSAA) might here show particular promise.

*What this study adds:* This is the first study to systematically assess the ability of both CSF NfL and CSF α-synuclein seeding activity to distinguish clinically diagnosed MSA (MSA_clin_) from SAOA, thereby offering a window into underlying MSA biology in patients *in vivo.* Our findings suggest that the rate of axonal degeneration is most pronounced in early MSA disease stages but decreases with longer disease duration; whereas α-synuclein seeding signal activity increases as MSA-related disease burden accumulates. Finally, it demonstrates the impact of a combined molecular fluid signature of MSA for improving trial design: a biomarker-based stratification of MSA subjects in future MSA treatment trials combining NfL plus α-synuclein seeding activity allows to reduce sample sizes by 20% compared to NfL alone.

*How this study might affect research, practice or policy:* The findings from this study may help to molecularly diagnose patients with MSA against overlapping and reciprocally mimicking conditions such as SAOA, in particular and even in early disease stages. Moreover, they might lay the foundation for a future biologically-informed diagnostic framework of MSA; support trial stratification for more efficient upcoming MSA treatment trials; and might facilitate molecular treatment effect monitoring in MSA, in particular in synuclein-targeted treatment trials.

## Introduction

Multiple system atrophy (MSA) is a rapidly progressive, adult-onset neurodegenerative disorder characterized by a combination of autonomic failure, cerebellar ataxia and/or parkinsonism, pathologically marked by glial cytoplasmic inclusions containing misfolded α-synuclein [1–6]. Among its closest clinical differentials is sporadic adult-onset ataxia (SAOA), a heterogeneous clinical category of late-adult-onset cerebellar ataxias not fulfilling the diagnostic criteria for MSA [7–10], and generally not considered to include synucleinopathy diseases [11]. Given their substantial clinical overlap, differentiating MSA from SAOA can be clinically challenging, particularly in early disease [12, 13], leading to potential diagnostic misclassification, suboptimal clinical management, and error-prone subject stratification for now emerging targeted MSA treatment trials [4, 6, 14].

Optimized clinical diagnostics and current neurodegenerative trials increasingly rely upon biological signatures of the respective disease to allow diagnosis and, if possible, initiate interventions in early disease stages prior to significant decline (see e.g. biological definitions of Alzheimer’s disease [15] and Parkinson’s disease [16, 17]). Identifying a mechanistically informed molecular biofluid signature of MSA might allow to optimize clinical diagnostics, *inter alia* by enabling its differentiation from SAOA even in early disease stages, and support trial stratification for more efficient upcoming MSA treatment trials. It should here ideally reflect also primary disease mechanisms, not only secondary degeneration processes [18]. Such a molecular signature might be enabled in MSA by combining a fluid marker of neuroaxonal degeneration with a mechanistic fluid marker closer to the primary disease mechanism. Specifically, neurofilament light chain (NfL)–elevated in MSA compared to controls and other ataxias [7, 9, 19–21]–presents a promising sensitive marker of neuroaxonal degeneration [22, 23] yet reflects an only rather non-specific downstream event in the pathophysiology cascade of MSA [5, 6]. In turn, α-synuclein seeding activity, measured in CSF using α-synuclein seed amplification assay (synSAA), might reflect a mechanistic fluid marker closer to the primary disease mechanism. Yet while sodium phosphate buffer (SPB) synSAA protocols (SBP-SAA) can consistently detect the seeding activity typical of Lewy body disease (LBD), they have shown limited ability to capture the distinct seeding activity characteristic of MSA [24, 25]. Optimized Piperazine-N,N’-bis(2-ethanesulfonic acid) (PIPES) synSAA protocols (PIPES-SAA) tailored for MSA have been recently developed [26–28]. Combining these two complementary markers of MSA biology, NfL reflecting neuroaxonal degeneration and synSAA reflecting more disease-specific α-synuclein seeding activity, may optimize clinical diagnostics of MSA, *inter alia* by enabling its differentiation from SAOA even in early disease stages. On a more general level, it might help to define a biologically informed CSF signature of MSA [29]. Such a biomarker-based MSA framework could improve diagnostic accuracy, enable molecularly-informed trial stratification, and support treatment-response monitoring in synuclein-targeted trials [30, 31].

Here, we assess whether CSF NfL and synSAA distinguish clinically diagnosed MSA (MSA_clin_) from SAOA and examine their diagnostic performance, disease-duration associations, and implications for biomarker-based stratification in future MSA trials.

## Materials and methods

### Cohort characteristics

Patients were recruited from the SPORTAX cohort, coordinated by the German Center for Neurodegenerative Diseases [9], and from the Ataxia Outpatient Clinic, Tuebingen. Diagnoses of MSA_clin_ were assessed using current consensus criteria: the MDS MSA diagnostic criteria [1] in 30 patients (n=15 clinically established, n=15 clinically probable MSA) and the second consensus criteria [32] in 2 patients (both probable MSA; 2022 MDS criteria not yet available at assessment). This study allowed inclusion of either MSA_clin_ subtype, i.e., parkinsonism-predominant (MSA-P) or cerebellar-predominant (MSA-C). MSA-C and MSA-P were both included in the primary analysis, as they represent transient clinical endophenotypes based on predominant motor symptoms at the time of evaluation, yet along a continuous spectrum of MSA as the single underlying disease process, with both extensive pathological overlap [33, 34] as well as strong clinical convergence over time [6, 35, 36]. The cohort was particularly enriched for MSA-C (n=22 MSA-C, n=10 MSA-P) given the study’s research aim and recruitment via SPORTAX and the Ataxia Outpatient Clinic. The remaining patients were classified as clinical SAOA (n=28), screened negative for genetic or secondary ataxia causes by the standard SPORTAX protocol (see Supplementary Methods). As clinical reference groups for benchmarking and interpretation of the PIPES-SAA results, we included, as positive controls, patients with LBD (LBD control_clin_, n=7) and, as negative controls, neurological ward controls (NWC_clin_, n=9). Additional cohort and CSF handling details are provided in the Supplementary Methods.

All patients provided written informed consent. The SPORTAX study is registered with ClinicalTrials.gov (NCT02701036), approved by local ethics committees, with additional approval for this analysis (353/2022BO2).

### NfL quantification

NfL concentrations were measured on an HD-X analyzer (Quanterix, USA) using the single molecule array (Simoa) N2PB kit (lot #504401). Details are provided in the Supplementary Methods.

### PIPES-SAA

The PIPES-SAA was performed using predefined fluorescence thresholds for sample result determination as previously described [28], with study-specific optimizations implemented in Tuebingen. Given recently highlighted challenges for inter-laboratory reproducibility of synSAA results [37], a technical validation of the PIPES-SAA was performed, using an independent cohort of clinically diagnosed LBD (LBD_clin_) and MSA_clin_ cases pre-screened for α-synuclein seeding. Details are provided in the Supplementary Methods.

### Statistical analysis

Normality and lognormality were assessed by visually inspecting histograms and Quantile-Quantile plots. Sex differences were assessed using Fisher’s exact test. Age at visit was compared using Welch’s t test; disease duration and global disability scale using log-normal Welch’s tests.

NfL values were log-transformed and age-adjusted using ordinary least squares (OLS) regression. Group comparisons of OLS age-adjusted residuals were assessed by the Mann–Whitney U test. Effect sizes were expressed as rank-biserial correlation (r). Multi-group comparisons used the Kruskal-Wallis test with Dunn’s post-hoc correction.

Diagnostic performance was assessed by receiver operating characteristic (ROC) analysis. Age-adjusted NfL values were derived using quantile regression (τ=0.5), subsequently back-transformed to the pg/mL scale, and thresholds defined by the Youden index within the study cohort to dichotomize NfL concentrations as above (N⁺) or below (N⁻) the threshold. SynSAA readouts were classified as positive (S⁺) or negative (S⁻) for synucleinopathy; inconclusive results, after a second independent measurement, were excluded. Sensitivity was defined as the percentage of MSA_clin_ classified as N⁺ or S⁺ and specificity as SAOA classified as N⁻ or S⁻; for the combined NfL+synSAA approach, sensitivity required both S⁺ and N⁺ in MSA_clin_, while specificity included SAOA with S⁻, N⁻, or both.

Associations between biomarker readouts and disease duration were assessed using Spearman rank correlation. Additional analyses focusing only on the MSA-C subgroup of MSA_clin_ were performed to assess consistency of the main diagnostic and disease duration findings independent of inclusion of MSA-P cases.

To estimate the effect of specificity of patient selection on required sample sizes in a hypothetical disease-modifying MSA treatment trial, we used as exemplary primary outcome the 24-month progression in UMSARS [35]. A fraction (1-specificity) of patients included was assumed to be SAOA. Detailed statistical methods, including modeling assumptions and effect size calculations, are described in the Supplementary Methods.

## Results

### Technical validation of the α-synuclein seed amplification assay

Given recently highlighted challenges for inter-laboratory reproducibility of synSAA results [37], a technical validation of the PIPES-SAA was performed prior to the main study using an independent cohort of LBD_clin_ (n=5), MSA_clin_ (n=10) and non-seeder neurological ward controls (NWC, n=4) cases. The assay discriminated synucleinopathy from NWC and differentiated LBD (type 1 seeds) from MSA (type 2 seeds), correctly classifying 100% of LBD_clin_ (5/5), 90% of MSA_clin_ (9/10) and 100% of NWC (4/4) cases, with distinct kinetic AUC values (1.1 × 10^6^ for LBD_clin_, 2.6 × 10^5^ for MSA_clin_, and 4.8 × 10^3^ for NWC at gain setting of 900) (**Supplementary Results and SFig. 1**). LBD showed the highest Fmax and AUC values, MSA intermediate signals, and controls only background noise. These results confirm the technical robustness and diagnostic validity of the PIPES-SAA in our lab, particularly for detecting the MSA-specific type 2 seeds.

### Characteristics of the main study cohort

Our study cohort included 32 patients with MSA_clin_ and 28 with SAOA, with demographic and clinical details summarized in **Table 1**.

**Table 1.** Demographic and clinical characteristics of the study cohort. Patients with MSAclin (n = 32) and SAOA (n = 28) were included. Values are n or mean ± SD. Group comparisons were performed as indicated in the Methods section, with p-values reported accordingly. MSAclin = clinically diagnosed multiple system atrophy; MSA-C = cerebellar-predominant MSAclin; MSA-P = parkinsonismpredominant MSAclin; SAOA = sporadic adult-onset ataxia; F/M = female/male; Global disability scale = UMSARS Part IV; CSF = cerebrospinal fluid; NfL = neurofilament light chain

| | MSA<sub>clin</sub><br>(n or mean ± SD) | SAOA<br>(n or mean ± SD) | p-value |
| --- | --- | --- | --- |
| **Total included** | 32 | 28 | — |
| **Sex (F/M)** | 15 / 17 | 12 / 16 | 0.799 |
| **Age at visit** | 59.3 ± 6.9 | 63.5 ± 12.3 | 0.117 |
| **Disease duration (years)** | 3.3 ± 1.9 | 8.2 ± 5.6 | <0.0001 |
| **Subtype** | 22/10 | — | — |
| **Global disability scale** | 2.7 ± 0.9 | 2.1 ± 1.0 | 0.008 |
| **CSF NfL (pg/mL)** | 3858.5 ± 2657.6 | 997.2 ± 750.1 | <0.0001 |

### CSF NfL discriminates MSA_clin_ from SAOA

Age-adjusted CSF NfL levels were significantly higher in MSA_clin_ (3859±2658 pg/mL) than in SAOA (997±750 pg/mL, p<0.0001, r=0.88) (**Fig. 1B**). ROC analysis yielded an AUC of 0.94 (95% CI 0.86–0.99, p<0.0001; **Fig. 1C**). The optimal Youden-derived age-adjusted threshold was 1384 pg/mL, which was subsequently used to dichotomize subjects into NfL above (N⁺) or below (N⁻) threshold (**Fig. 1A**). Accordingly, 31/32 MSA_clin_ cases were classified as N⁺, while 24/28 SAOA cases were classified as N⁻ (**Fig. 1D**). With clinical MSA diagnosis according to MDS criteria as reference, diagnostic performance was: sensitivity 96.9% (95% CI 83.8–99.9), specificity 85.7% (95% CI 67.3–96.0), with PPV 81.9% (95% CI 69.3–95.1) and NPV 97.6% (95% CI 92.3–100.0) (**Fig. 1E**). These diagnostic performance values of NfL were confirmed also when comparing only the MSA-C subgroup to SAOA (**SFig. 2A-B**). For reference, CSF NfL levels were also assessed in LBD control_clin_ and NWC_clin_. Age-adjusted levels were higher in MSA_clin_ than NWC_clin_ (p=0.009) and LBD control_clin_ (p=0.04), both showing lower values in ranges comparable to SAOA (**SFig. 3**).

**Fig. 1.**
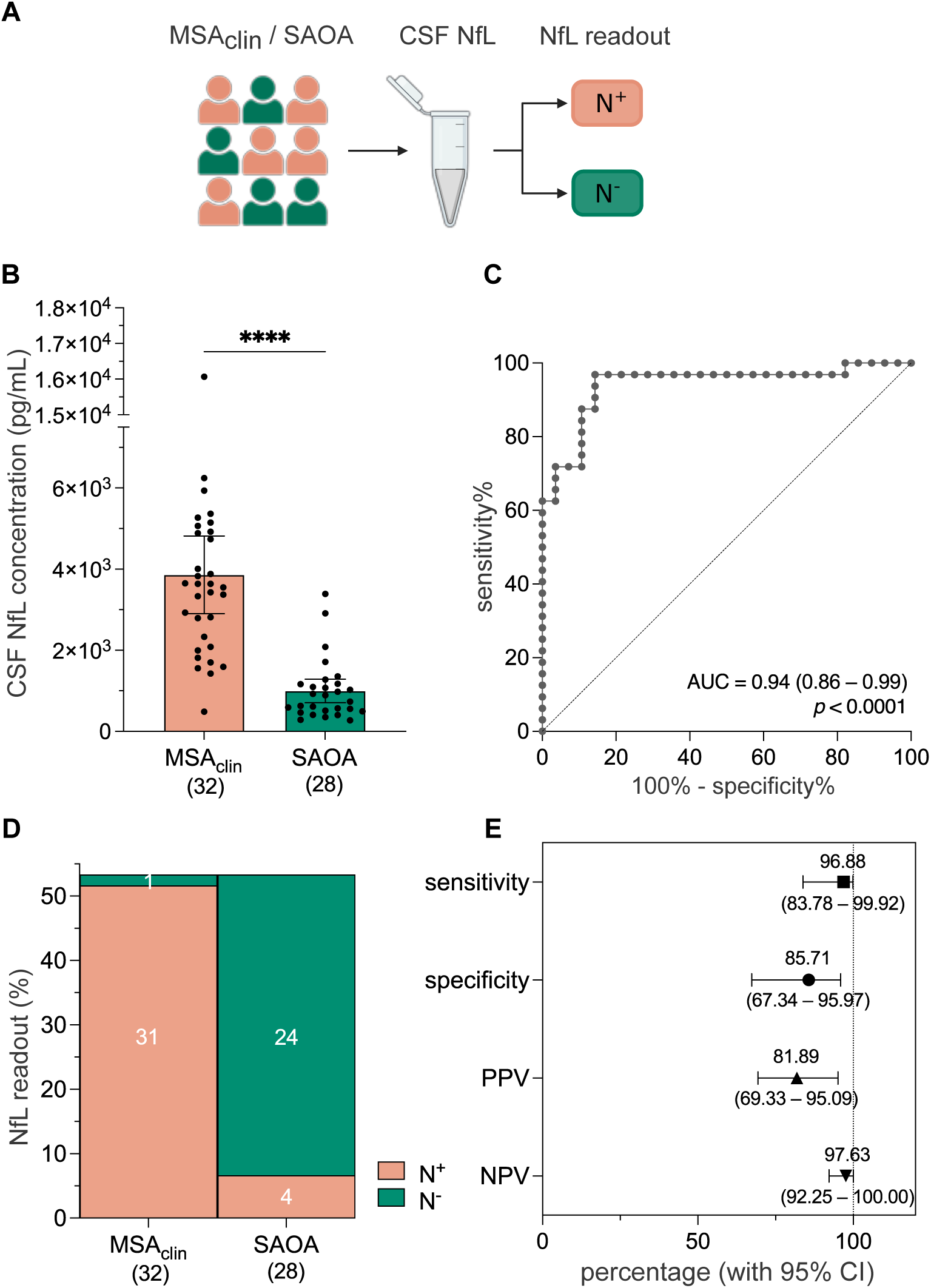
CSF NfL discriminates MSA_clin_ from SAOA. **(A)** Schematic overview of the analysis design: CSF neurofilament light chain (NfL) concentrations were measured in MSA_clin_ (n = 32) and SAOA (n = 28), and cases were dichotomized into NfL-high (N⁺) and NfL-low (N⁻) categories. (**B)** Bar plot of raw CSF NfL concentrations in MSA_clin_ and SAOA, with each datapoint representing the mean of two technical replicates per case. Bars indicate group means ± 95% CI. Group comparison was performed on age-adjusted residuals using the Mann–Whitney U test (U = 53, ***p < 0.0001, r = 0.88). (**C)** Receiver operating characteristic (ROC) curve illustrating separation between MSA_clin_ and SAOA; threshold (1383.56 pg/mL) was determined by the Youden index on age-adjusted residuals. (**D)** Marimekko plot summarizing the classification of MSA_clin_ (n = 32) and SAOA (n = 28) cases by NfL readout. Readouts were categorized as N⁺ or N⁻. The y-axis indicates the percentage of cases in each category, with the number of cases shown in white within each stacked bar. (**E)** Forest plot of diagnostic performance metrics for distinguishing MSA_clin_ from SAOA, with point estimates and 95% CI shown. CI = confidence interval; NPV = negative predictive value; PPV = positive predictive value

### CSF PIPES-SAA discriminates MSA_clin_ from SAOA

CSF PIPES-SAA maximum fluorescence intensity (Fmax) values were consistently higher in MSA_clin_ compared to SAOA (**SFig. 4**). MSA_clin_ showed the characteristic intermediate Fmax values, namely distinctively lower than LBD control_clin_ (=type 1 seeds), but higher than the minimal Fmax values in NWC_clin._ In contrast, Fmax values in SAOA were indistinguishable from those of NWC_clin_ (**SFig. 4**).

Group-level synucleinopathy classification results were further analyzed, summarizing the proportions of synSAA-positive (S⁺) and synSAA-negative (S⁻) subjects for MSA_clin_ and SAOA (**Fig. 2A**), and differentiating these by type 1 and type 2 seeds. Seed-type distributions across all diagnostic groups are shown in **Fig. 2B**. Among MSA_clin_ (n=32), 78% were S⁺ (n=25), predominantly exhibiting type 2 seeds (n=23, 72%), with a single subject showing type 1 (n=1) or unresolved seed type (S⁺ undetermined, n=1), while 22% were S⁻ (n=7) (**Fig. 2B-C**). In contrast, 82% of all SAOA (n=28) were S⁻ (n=23), with only 14% S⁺ (n=4); here with 3/4 showing type 1 seeds and only 1/4 showing type 2 seeds (=4% of all SAOA). One sample yielded inconclusive results (n=1) (**Fig. 2B-C**). In the reference groups, LBD control_clin_ subjects were predominantly S^+^ with type 1 seeds (5/7, 71%), and the rest were S⁻ (2/7, 29%), while all NWC_clin_ were S^−^ (n=9; 100%) (**Fig. 2B**), thus confirming the overall assay performance in this study. The PIPES-SAA assay differentiated MSA_clin_ from SAOA with 78.1% sensitivity (95% CI 60.0–90.7), 85.2% specificity (95% CI 66.3–95.8), 77.9% PPV (95% CI 62.8–93.8), and 85.4% NPV (95% CI 77.2–93.7) (**Fig. 2D**). These diagnostic performance values of PIPES-SAA were confirmed also for distinguishing only the MSA-C subgroup from SAOA (**SFig. 2C-D**). Thus, overall, the PIPES-SAA identified a majority of MSA_clin_ cases with high specificity against SAOA.

**Fig. 2.**
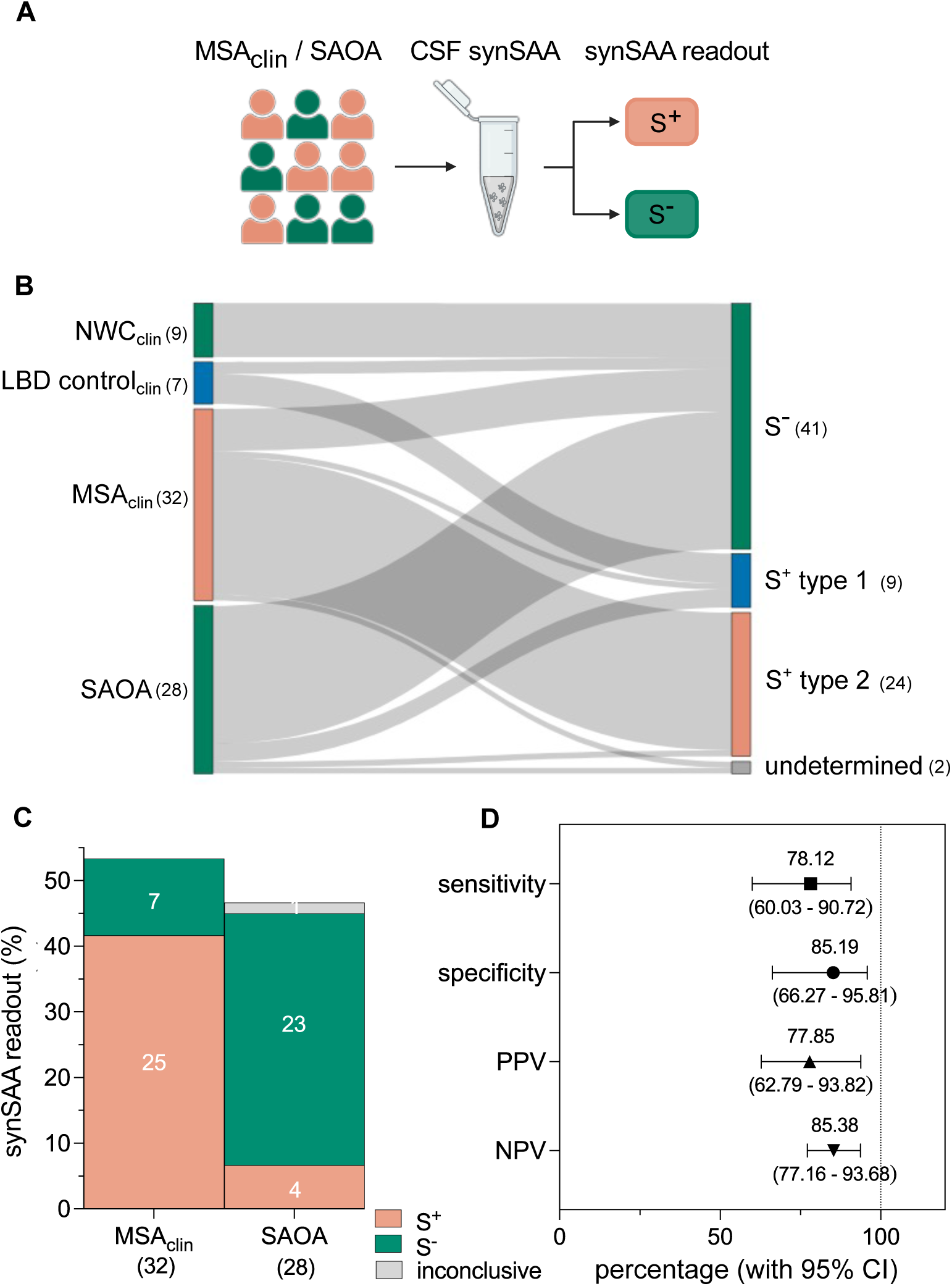
CSF PIPES-SAA discriminates MSA_clin_ from SAOA. (**A)** Schematic of the analysis design: CSF PIPES-SAA was measured in MSA_clin_ (n = 32) and SAOA (n = 28), with readouts classified as positive (S⁺), negative (S⁻), or inconclusive. (**B)** Sankey plot illustrating group-wise distribution of synSAA readouts. Each flow originates from the diagnostic group on the left (MSA_clin_, SAOA, and the clinical reference groups LBD control_clin_ and neurological ward controls [NWC_clin_]) and connects to the synSAA readout categories on the right (S⁻ or S⁺ further subclassified based on their seeding pattern as either type 1 or type 2, defined by distinct kinetic fluorescence profiles, or undetermined in case of inconclusive seed presence or type). Line thickness reflects the number of cases, with case counts shown in parentheses. Colors indicate the readout categories (right): green = S⁻ (negative), orange = S⁺ type 2, blue = S⁺ type 1, and grey = undetermined. Diagnostic groups (left) are shaded according to their typical expected readout for visualization. (**C)** Marimekko plot summarizing the classification of MSA_clin_ (n = 32) and SAOA (n = 28) cases by synSAA readout. Readouts were categorized as positive (S⁺), negative (S⁻), or inconclusive. The y-axis indicates the percentage of cases in each category, with case counts shown in white within each stacked bar. (**D)** Forest plot of diagnostic performance metrics for distinguishing MSA_clin_ from SAOA, showing point estimates and 95% CI. synSAA = α-synuclein seed amplification assay; CI = confidence interval; NPV = negative predictive value; PPV = positive predictive value

### Divergent dynamics of NfL and synSAA across disease duration stages in MSA_clin_

To explore biomarker dynamics across disease duration in MSA_clin_, we analyzed associations of CSF NfL and synSAA readouts with disease duration in a cross-sectional design. Age-adjusted NfL levels were elevated early but showed lower values with longer disease duration (n=32, Spearman r=-0.45, p=0.01) (**Fig. 3A**). This negative correlation retained its trend after excluding Cook’s distance outliers (n=28, r=-0.34, p=0.079, data not shown). Next, disease duration was dichotomized according to recent clinical trial protocols into early (<4 years from onset, n=22) and late (≥4 years, n=10) stages, revealing statistically significant lower NfL levels in late-stage compared to early-stage MSA_clin_ (p=0.01, r=0.55) (**Fig. 3B**). In contrast, PIPES-SAA Fmax values where higher with advancing disease (Spearman r=0.42, p=0.02) (**Fig. 3C**). Its positive correlation also remained significant after excluding Cook’s distance outliers (n=28, r=0.39, p=0.042, data not shown). A sensitivity analysis restricted to MSA_clin_ samples exhibiting type 2 seeds confirmed a consistent and statistically significant positive association (n=24, p=0.02, r=0.47, data not shown). The late-stage MSA_clin_ group showed significantly higher Fmax values compared to the early-stage group (p=0.01, r=0.55) (**Fig. 3D**). Analyzing only the MSA-C subgroup (n=22) confirmed both, a negative correlation of disease duration with NfL (Spearman r=-0.26, p=0.25) (**SFig. 5A**), and a positive correlation of disease duration with synSAA Fmax (Spearman r=0.48, p=0.02) (**SFig. 5B**). Together, these findings indicate divergent trajectories of NfL and synSAA in MSA_clin_: NfL, reflecting neuroaxonal injury, peaks early and declines over time; whereas synSAA Fmax, indicating α-synuclein seeding activity, increases with longer disease duration.

**Fig. 3.**
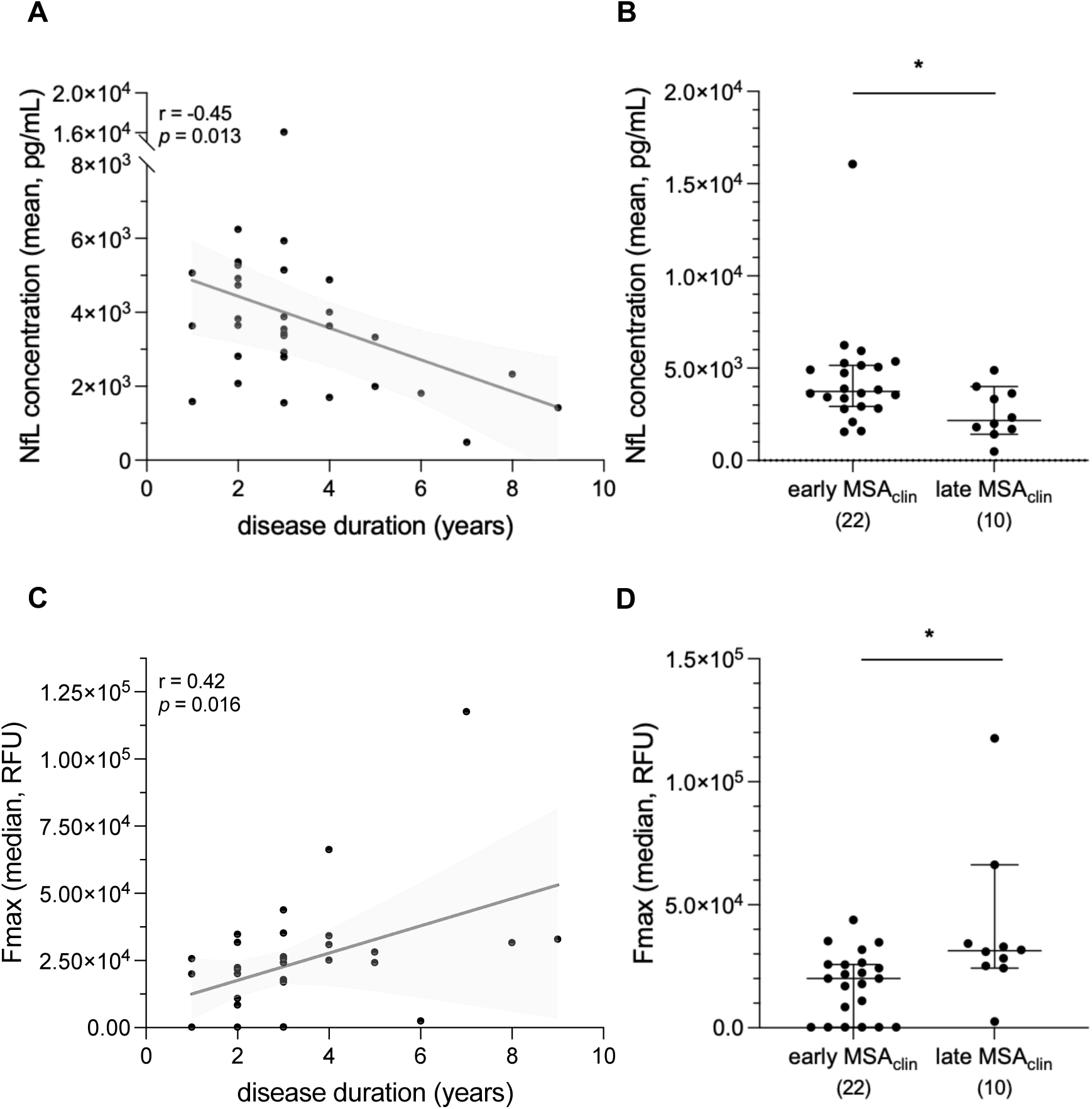
NfL and synSAA show divergent associations with disease duration in MSA_clin_. **(A)** Scatter plot of CSF NfL versus disease duration in MSA_clin_ (n = 32). Each datapoint represents the raw mean NfL concentration of technical duplicates per case. Line (black) shows linear fit and its 95% CI (grey) for visualization. The association was tested using Spearman correlation on age-adjusted residuals (r = – 0.45, p = 0.013). **(B)** Scatter dot plots comparing mean NfL concentrations between early- (<4 years from onset, n = 22) and late- (≥4 years, n = 10) stage MSA_clin_ cases. Plots show the median (line) with 95% CI (error bars); all individual cases are shown as datapoints. Group differences assessed using Mann-Whitney U test on age-adjusted residuals (U = 50, *p < 0.05, r = 0.55). **(C)** Scatter plot of PIPES-SAA maximum fluorescence intensity (Fmax, RFU) versus disease duration in MSA_clin_ cases (n = 32). Each datapoint represents the median Fmax of three technical replicates per case. Line (black) shows linear fit with 95% CI (grey) for visualization; association tested by Spearman correlation (r = 0.42, p = 0.016). **(D)** Scatter dot plots comparing median Fmax between early- and late-stage MSA_clin_ subjects. Plots show the median with 95% CI; each case indicates one datapoint. Group differences were assessed by Mann-Whitney U test (U = 49, r = 0.55). synSAA = α-synuclein seed amplification assay; NfL = neurofilament light chain; CI = confidence interval; RFU = relative fluorescence units

### Combining NfL with synSAA reduces required sample sizes in treatment trials compared to NfL alone

As biomarker-based subject stratification can markedly reduce required sample sizes in clinical trials [30, 31, 38–40], we estimated and compared sample size requirements for hypothetical MSA treatment trials based on biomarker-based stratification, taking as primary outcome UMSARS progression (21.9±11.9 points/24 months) [35]. Estimates were derived based on the diagnostic specificities either of NfL alone or in combination with synSAA (NfL+synSAA) scaled by the literature-reported specificity of the clinically probable MDS MSA criteria (0.74) [41] to approximate the diagnostic performance expected relative to pathology-confirmed MSA and yield realistic trial sample size estimates true to disease. Assuming a 50% therapeutic effect, stratification by elevated age-adjusted NfL (specificity=85.7%) required 161 participants per arm (**SFig. 6**). We next examined whether NfL+synSAA further improved trial efficiency. The NfL+synSAA classifier–defined as cases positive for both SAA and elevated age-adjusted NfL–achieved 96.3% specificity (95% CI 81.03–99.91) and 75.0% sensitivity (95% CI 56.6–88.5). Incorporating both biomarkers (NfL+synSAA) reduced the required sample size by 20%, to 129 participants per arm at 50% therapeutic effect (**SFig. 6**).

## Discussion

In this study, we evaluated the diagnostic and biological utility of two complementary fluid biomarkers–α-synuclein seeding activity and NfL–for improving the discrimination of MSA_clin_ from SAOA, a key phenotypic MSA-mimic. Moving beyond comparisons with healthy or general neurological controls, we addressed the real-world challenge of distinguishing MSA from its main phenotypically overlapping syndromes [8,11–13,35] by example of SAOA. SAOA often presents with features resembling MSA-C [8, 12–14], and diagnostic discrimination is difficult particularly in early disease (<4 years) [42]. The primary analyses of this study were performed in the full MSA_clin_ cohort, since MSA-C and MSA-P represent transient clinical endophenotypes based on predominant motor symptoms at the respective time of evaluation, along a continuous spectrum of MSA as a single underlying disease process, with both extensive pathological overlap [33, 34] as well as strong clinical convergence over time [6, 35, 36]. Correspondingly, subgroup analyses of only the MSA-C subcohort confirmed all main findings. Our findings highlight how combining a sensitive marker of neuroaxonal degeneration (NfL) with a mechanistically specific marker of synucleinopathy (synSAA) can enhance diagnostic precision and support a biologically grounded framework for MSA.

### NfL discriminates MSA_clin_ from SAOA as a biomarker of axonal degeneration

CSF NfL levels showed high discrimination between MSA_clin_ and SAOA (AUC=0.94). Its high sensitivity (97%) and NPV (98%) indicate that low NfL level (N^−^) makes MSA_clin_ unlikely. This high discriminatory performance of CSF NfL might support both clinical management and trial inclusion as demonstrated here for already quite early stage MSA_clin_ patients (<4 years of disease) where diagnostic discrimination on clinical grounds alone is particularly difficult, but where early diagnosis is of particular importance for patient life planning and early stratification into trial before disease is too advanced. These findings corroborate and extend previous studies demonstrating elevated NfL levels in MSA relative to SAOA [7, 9, 19].

Interestingly, the sensitivity, specificity, and proposed cut-off observed in our study to discriminate MSA_clin_ from SAOA (97%, 86%, 1384 pg/mL) were similar to those reported for discriminating MSA from LBD (97%, 90%, 1400 pg/mL, [43]), although based on a different assay platform (ELISA-based) [43]. Taken together, these findings provide convergent evidence supporting a CSF NfL cut-off in the range of 1384-1400 pg/mL for discriminating MSA both from non-MSA overlap conditions (LBD, SAOA) and across different assay platforms (SIMOA, ELISA).

While specificity of the CSF NfL cut-off was high (86%), it was not as high as sensitivity, with a substantial share of SAOA cases (14%) showing N^+^. This demonstrates that the rate of axonal degeneration can be substantially increased also in patients from the biologically very heterogeneous group of SAOA [7]. Thus, NfL improves classification of MSA_clin_ by complementing clinical diagnosis with an objective measure of neuroaxonal degeneration, but its limited specificity highlights the need for more mechanistically specific biomarkers.

### α-synuclein seeding activity distinguishes MSA_clin_ from SAOA as a mechanistic biomarker

SynSAA directly detects α-synuclein seeding activity, a defining pathological hallmark of MSA. Our PIPES-SAA protocol identified the majority (25/32) of MSA_clin_ cases and was negative in most SAOA (23/28), demonstrating high discrimination to distinguish MSA_clin_ from SAOA. Its relatively high specificity (86%) supports selective identification of synucleinopathy when S^+^. Its sensitivity was moderate (78%), indicating that some MSA_clin_ remain S^−^. Notably, these S^−^ cases were observed in both MSA-C (4/22, 18%) and MSA-P (3/10, 30%). The rather only moderate sensitivity, i.e., a substantial share of S^−^ MSA_clin_ patients (7/32=22%) may reflect: (i) a subcohort of S^−^ MSA patients with a different molecular/pathological disease evolution than S^+^ MSA patients, analogous to S^+^ vs S^−^ PD or DLB disease subcohorts [44–47]; (ii) early disease stages, with seeding activity still below synSAA detection limits; (iii) general current assay detection limitations; or (iv) patients falsely clinically classified as MSA even by the latest MDS MSA diagnostic criteria, as up to 12% of sporadic adult-onset ataxia meeting MSA_clin_ criteria have been shown to carry an alternative, non-MSA disease cause [7]. Conversely, the 15% S^+^ SAOA cases may reflect early MSA patients not yet meeting the MDS MSA diagnostic criteria or, alternatively, incidental α-synuclein age-related Lewy body co-pathology [48].

SynSAA is mechanistically specific for α-synuclein seeding activity, in contrast to NfL, which reflects non-specific neuroaxonal damage. Thus, while synSAA alone does not outperform NfL statistically, an S^+^ versus S^−^ result might provide mechanistic diagnostic information for clinical diagnosis or trial inclusion/exclusion. For example, adding synSAA to NfL may support biologically informed diagnostic frameworks, when differentiating MSA from SAOA remains uncertain. For instance, one MSA_clin_ case showed exceptionally high NfL (Fig. 1B, highest datapoint), confirmed by two independent NfL assessments, but yielded an S^−^ result. Although formally included as MSA_clin_, based on the–largely clinical–MDS MSA criteria, this discordant biomarker profile may indicate an underlying alternative rapidly progressive neurodegenerative disease (e.g., prion disease) not diagnosed. Such cases suggest that combined NfL and synSAA assessment may prompt diagnostic reevaluation in atypical presentations.

### Increase of synSAA Fmax with disease duration, and complementary roles of synSAA and NfL in MSA_clin_

The median synSAA Fmax values were higher with advanced disease duration, suggesting increasing synSAA signal as disease burden accumulates in MSA_clin_. If confirmed longitudinally, this might indicate its potential as a mechanistically linked progression biomarker in MSA, analogous to type 1 synSAA readouts in longitudinal PD studies [46, 49–51]. More broadly, these findings suggest that the synSAA might provide both *trait* and *state* information in MSA_clin_: while synSAA positivity (S^+^/S^−^) functions as a dichotomous trait marker indicating the presence or absence of synucleinopathy, quantitative synSAA metrics (such as Fmax) may capture graded disease states. If confirmed in longitudinal and neuropathology-enriched studies, this finding would thus extend approaches on possible dual information derived from synSAA in LBD [51–53] to, for the first time, synSAA in MSA_clin_.

In contrast to synSAA Fmax, NfL levels were highest in early MSA_clin_ disease, but lower in patients with longer duration, consistent with other reports [9, 54, 55]. If confirmed by longitudinal studies, this finding indicates that the rate of neuroaxonal decline might be high earlier in the disease course and declining thereafter, when substantial disease-related neuronal loss has already occurred. The divergent trajectories of synSAA and NfL might suggest different aspects of the underlying disease processes relative to disease stage/duration: while NfL is sensitive to the rate of neuroaxonal decline, which is highest in MSA_clin_ in early stages (as shown here and elsewhere [9, 54, 55], synSAA might reflect an accumulating tendency of synuclein seeding signals–which increase with disease duration. If confirmed by required longitudinal studies, such divergent associations of NfL and synSAA with disease duration could thus indicate that these biomarkers might capture complementary aspects of underlying MSA biology relative to disease duration from *living* patients.

### Toward a biological definition of MSA

Therapeutic trials in neurodegenerative diseases increasingly adopt biological definitions to enable earlier intervention and improve diagnostic precision for enrolling the appropriate target population. While such biological definitions–often integrating fluid biomarkers–have been proposed for AD [56] and PD [17], they remain less well established for MSA [29]. Our findings provide preliminary evidence supporting such a framework in MSA by identifying two complementary biomarkers, NfL and synSAA. Biomarkers informing such a biological framework should ideally (i) reflect not only secondary degeneration processes, but also primary disease mechanisms; and (ii) with increasing, rather than decreasing, differences relative to healthy controls with advancing disease duration [18]. This might indeed be enabled for MSA by: NfL as a non-specific marker reflecting neuroaxonal degeneration, combined with synSAA as a mechanistic marker capturing MSA-specific α-synuclein aggregation processes–even distinct from those in PD (SFig. 1). Importantly, certain metrics of synSAA (Fmax) also increase, rather than decrease (like NfL), relative to healthy controls with advancing disease duration. Recent regulatory precedents support the translational viability of combined biomarker signatures, such as e.g. the FDA-cleared Lumipulse G plasma assay for pTau217 and Aβ42 in AD [57].

If these findings are confirmed by post-mortem correlation and longitudinal CSF studies, they might pave the way toward a biological definition of MSA (“S^+^N^+^ framework”). The MDS clinical diagnostic framework for MSA has defined a category *clinically probable* designed to “enhance sensitivity while maintaining specificity”; and a category *clinically established* aiming for “maximum specificity with acceptable sensitivity” [1, 41]. By analogy, our biomarker findings suggest a similar balance between sensitivity and specificity: elevated NfL alone (high sensitivity, moderate specificity) may correspond to a category *biomarker-probable*; whereas concordant NfL+synSAA positivity (higher specificity, moderate sensitivity) may correspond to a category *biomarker-established*. While attractive, this analogy for now remains conceptual, and will require thorough neuropathological and longitudinal validation.

### Rationale for, and impact on, biomarker-based enrichment trials

Biomarker-based participant enrichment can improve the design and interpretability of MSA treatment trials, where diagnostic heterogeneity and inclusion of mimics such as SAOA can dilute measurable treatment effects [30, 31]. Trial-size modeling based on UMSARS progression data indicated that the higher specificity of NfL+synSAA can reduce required sample sizes by ∼20% compared with NfL alone. By increasing specificity, the combined NfL+synSAA approach limits enrollment of non-MSA participants into MSA trials, thereby enhancing statistical power, reducing costs, and strengthening the ethical justification for exposing participants to investigational therapies. Although less sensitive than NfL alone, the NfL+synSAA approach defines a high-confidence, biologically characterized subgroup and may improve efficiency of synuclein-targeted trials by ensuring target engagement.

### Limitations

Despite high internal consistency, external replication across independent sites and plate-reader systems will be essential to confirm generalizability. This study is further limited by the relatively small sample size, the absence of additional complementary biomarkers (e.g., imaging or hyposmia assessments), and in particular its cross-sectional nature; accordingly, interpretations of biomarker dynamics over disease course should be considered exploratory, warranting confirmation by future longitudinal studies. Moreover, clinical diagnosis served as the reference standard as neuropathological confirmation was unavailable. Confirmation by future studies in pathologically confirmed MSA cohorts as gold standard is thus warranted. Yet, application of the MDS MSA criteria used here for clinically probable MSA shows 95% sensitivity and 94% specificity against postmortem neuropathological diagnosis [42], thus serving as a valuable proxy for the actual gold standard, supporting the interpretability of the present findings. In sum, current results should be considered hypothesis-generating and warrant further validation in post-mortem and prospective longitudinal cohorts.

### Future directions

This study provides a proof-of-concept for integrating CSF NfL and synSAA to biologically define and stage MSA against SAOA. Multicenter, longitudinal, and pathology-validated studies will be important to confirm reproducibility, establish clinical-grade cut-offs, and verify stage-dependent biomarker trajectories at the single-patient level. Translation of this framework to blood-based formats could extend diagnostic accessibility. Upon validation, such a framework could enable biologically informed diagnostics, support recruitment, and facilitate treatment-response monitoring in MSA–particularly in synuclein-targeted treatment trials.

## Statements and Declarations

### Conflicts of interest

VK has declared that no conflict of interest exists.

LCM has declared that no conflict of interest exists. LC-M is an employee of Amprion, a biotech company dedicated to the commercialization of seed amplification assay technologies. LCM is inventor of several patents related to seed amplification assay technologies, which have been assigned to Amprion.

LB has declared that no conflict of interest exists.

AH have declared that no conflict of interest exists.

TK has declared that no conflict of interest exists.

JF has received consultancy honoraria from Vico therapeutics and Biogen, unrelated to the present manuscript. KB has declared that no conflict of interest exists.

LS has declared that no conflict of interest exists.

BR has declared that no conflict of interest exists.

DM has declared that no conflict of interest exists.

MS has received consultancy honoraria from Ionis, UCB, Prevail, Orphazyme, Biogen, Insmed, Servier, Reata, GenOrph, AviadoBio, Biohaven, Zevra, Lilly, Quince, and Solaxa, all unrelated to the present manuscript.

### Funding

This work was supported, in part, by the Clinician Scientist program “PRECISE.net 2.0” funded by the Else Kröner-Fresenius-Stiftung (to L.B, L.S, D.M, M.S.); and by the project European Rare Disease Research Alliance (ERDERA), GA n°101156595, funded under call HORIZON-HLTH-2023-DISEASE-07 (to M.S.) by the European Union. D.M. was supported by the Clinician Scientist program of the Medical Faculty Tübingen (459-0-0) and the Elite Program for Postdoctoral researchers of the Baden-Württemberg-Foundation (1.16101.21). JF is funded within the Advanced Clinician Scientist Programme (ACCENT, funded by the Federal Ministry of Research, Technology and Space (BMFTR) under the funding code (FKZ): 01EO2107) and as a PI of the iBehave Network, sponsored by the Ministry of Culture and Science of the State of North Rhine-Westphalia and received funding from the National Ataxia Foundation (NAF). JF is a member of the of the European Reference Network for Rare Neurological Diseases. B.R. was supported by the Clinician Scientist program of the Medical Faculty Tübingen (478-0-0).

### Author contributions

Study conception: DM and MS; experimental design: VK, DM and MS; data acquisition and analyses: VK, LCM, LB and AH; diagnosis and clinical data acquisition: TK, JF, KB, LS, BR, DM and MS; data interpretation: VK, DM and MS; manuscript first draft: VK, DM and MS; manuscript review and editing: all authors; final manuscript approval: all authors.

### Generative AI Acknowledgment

We used ChatGPT (OpenAI; model GPT-5) for language polishing of early drafts. The authors verified and edited all content.

### Informed consent statement

Informed consent was obtained from all patients involved this study wherever necessary.

### Data availability statement

All data supporting the findings of this study are available within this paper and its supplementary information. Further information can be provided upon reasonable request to the corresponding author.

## Acknowledgements

We are grateful to the Biobanks at the Hertie Institute for Clinical Brain Research, Tuebingen (Claudia Schulte, Christian Deuschle) and at the German Center for Neurodegenerative Diseases (DZNE) Bonn, Bonn, Germany for providing CSF samples.

## Notes

### Author Declarations

The SPORTAX study was approved by the responsible ethics committees and registered with ClinicalTrials.gov (NCT02701036). The Ethics Committee of the University of Tuebingen gave additional ethical approval for this work (353/2022BO2).

