## Supplemental material for "A molecular biofluid signature of multiple system atrophy (MSA): CSF neurofilament light chain and α-synuclein seeding as complementary biomarkers allow to distinguish MSA from sporadic adult-onset ataxia"

### SUPPLEMENTARY MATERIAL

### Supplementary Methods

#### Cohort characteristics

Patients were recruited from the SPORTAX cohort, a registry-based, prospective, longitudinal, European multicenter observational study coordinated by the German Center for Neurodegenerative Diseases (DZNE) [1], and from the Ataxia Outpatient Clinic, Tuebingen. Inclusion criteria were defined to specifically identify cases of sporadic late-adult-onset degenerative ataxia: (i) age at onset/first visit older than age 30 years (resulting in a cohort with 99% of all patients with age at onset ≥ 40 years. (ii) informative and negative family history (no similar disorders in first- and second-degree relatives; parents older than 50 years, or, if not alive, age at death of more than 50 years, no consanguinity of parents); (iii) negative molecular genetic testing for Friedreich's ataxia (FRDA), spinocerebellar ataxia type 1 (SCA1), SCA2, SCA3, SCA6, and Fragile X), and (iv) absence of an identifiable acquired cause of ataxia, as defined by the following (in line with Giordano et al. 2017): (a) clinical: no onset of ataxia associated with stroke, encephalitis, sepsis, hyperthermia or heat stroke, chronic diarrhea, unexplained visual loss, alcohol abuse, chronic intake of anticonvulsant drugs, other toxic causes, malignancies; rapid progression defined as development of severe ataxia in less than 12 weeks; no insulin-dependent diabetes; (b) imaging: no evidence of multiple sclerosis, ischemia, hemorrhage, or posterior fossa tumor; no signal abnormalities on T2/FLAIR images, except those compatible with MSA-C; (c) laboratory: antineuronal antibodies negative (only required if disease duration less than 3 years); normal vitamin B12 levels; negative VDRL; normal thyroid function [2].

Based on these criteria, a total of 60 patients with available CSF were recruited. Patients were classified with clinical multiple system atrophy ($MSA_{clin}$) if meeting the diagnostic criteria at least for "clinically probable multiple system atrophy" (n=32 total), either according to the MDS MSA diagnostic criteria [3] (n=30 patients; thereof n=15 fulfilling criteria for clinically established and n=15 for clinically probable

MSA) or, in patients assessed before the availability of these latest criteria, according to the second consensus MSA diagnostic criteria [4] (n=2 patients, both probable MSA). This study allowed inclusion of either $MSA_{clin}$ subtype, i.e., parkinsonism-predominant (MSA-P) or cerebellar-predominant (MSA-C). MSA-C and MSA-P were considered jointly in the primary analyses as they represent transient clinical endophenotypes based on predominant motor symptoms at the respective time of evaluation, yet along a continuous spectrum of MSA as the single underlying disease process, with both extensive pathological overlap [5, 6] as well as strong clinical convergence over time [7-9]. The cohort was particularly enriched for MSA-C patients (n=22 MSA-C, n=10 MSA-P patients) given the study's research aim and its recruitment structure via the SPORTAX cohort and the Ataxia Outpatient Clinic, Tuebingen. The remaining patients were classified as clinical sporadic adult-onset ataxia (SAOA, n=28). Clinical assessments included the Unified Multiple System Atrophy Rating Scale (UMSARS) Part IV (global disability scale) [10].

As clinical reference groups for benchmarking and interpretation of the α-synuclein seed amplification assay (synSAA) using PIPES buffer (PIPES-SAA) results, we additionally included, as positive controls, patients with Lewy body disease (LBD $control_{clin}$, n=7) and, as negative controls, neurological ward controls ($NWC_{clin}$, n=9) into the study cohort. LBD $control_{clin}$ group comprised patients clinically diagnosed with either Parkinson's disease (PD, based on the UK Brain Bank Society criteria [11], n=1) or dementia with Lewy bodies (DLB; based on the fourth consensus report of the DLB consortium [12], n=6). The $NWC_{clin}$ group comprised patients clinically diagnosed with neurological diseases other than synucleinopathy disorders, e.g., polyneuropathy or idiopathic intracranial hypertension. All subjects of these clinical reference groups, except for one LBD $control_{clin}$ subject, were also previously reported and characterized in another study [13].

Given the recently highlighted challenges for the reproducibility of synSAA results across different laboratories [14], a technical validation of the PIPES-SAA was performed prior to the main study. For this, we included independent cohort of LBD and MSA cases that had been pre-screened for α-synuclein seeding in a previous study and were used here as experimental controls. Specifically, they comprised of $MSA_{clin}$

(n=10; 6 MSA-P and 4 MSA-C), LBD ($LBD_{clin}$, n=5; 4 DLB and 1 PD), and neurological ward controls (NWC, n = 4; 2 polyneuropathy and 2 idiopathic intracranial hypertension). Diagnostic criteria were applied as described for the main study cohort.

All patients provided written informed consent for this study at their respective centers. The SPORTAX study is registered with ClinicalTrials.gov (NCT02701036), approved by the ethics committee of University Tübingen (598/2011BO1), with additional approval for this analysis (353/2022BO2).

**CSF handling**

CSF samples were collected by lumbar puncture according to standardized procedures from the biobank facilities at the Hertie Institute for Clinical Brain Research, Tuebingen, Germany and the DZNE in Bonn, Germany. Samples were centrifuged at 2000 x *g* for 10 min at room temperature within 30 min of withdrawal, then pseudonymized, aliquoted into 0.3-0.5 mL volumes, and frozen at -80°C within 90 min after collection.

For this study, selected samples were thawed on ice and re-aliquoted into single-use aliquots of 140 µL (at least 2 per case, wherever possible) and one 30 µL aliquot, transferred to LoBind tubes (Eppendorf, Hamburg, Germany), and re-stored at –80°C, and thawed only once again at the time of measurement.

**NfL quantification**

NfL concentrations were measured on an HD-X analyzer (Quanterix, Billerica, MA, USA) using the ultrasensitive single molecule array (Simoa) Neurology 2-Plex B Advantage kit (N2PB, Quanterix, Billerica, MA, USA, lot #504401), according to the manufacturer's instructions. Prior to measurement, CSF was centrifuged at 10,000 × g for 5 min at 4°C and diluted in kit sample buffer. Measurements were run in technical duplicates. The assay's lower limit of quantification (LLoQ) was defined as the lowest standard meeting two conditions: (i) its signal was greater than the average signal for the blank plus nine times its standard deviation (SD), and (ii) it allowed a percentage recovery ≥ 100±20%, and was multiplied by the applied dilution factor of 40. The LLoQ across all measurements was 18.28 pg/mL. Sample measurements were included only if duplicate average enzyme per bead (AEB) coefficients of variation (CVs) were <20%. Across all samples, the mean AEB CV was 4.3%±3.9% (range 0.004–15.6%). The measurements were performed blinded to clinical diagnosis.

**PIPES-SAA**

The PIPES-SAA was performed as previously described by Amprion [13] with adaptations optimized for implementing the assay in Tuebingen for this study. As published earlier [13], predefined Fmax thresholds were used to distinguish type 1 and type 2 α-synuclein seeds and to classify samples as synSAA-positive ($S^+$) or synSAA-negative ($S^–$). Specifically, the original 3.2 mm $Si_3N_4$ beads were replaced with 3 mm glass beads (Sigma-Aldrich, St. Louis, MO, USA; Cat# 104015) due to discontinuation of the previously used type by the manufacturer. For each synSAA run, CSF samples were thawed in cold tap water immediately prior to use. Aliquots were used once and were not subjected to repeated freeze–thaw cycles. Briefly, two 3 mm glass beads (Sigma Aldrich, St. Louis, MO, USA; Cat#104015, lot#34821999) were placed into each well, either using sterile forceps or an Amprion-designed bead dispenser. A total of 40 µL CSF per sample was loaded in triplicates into black clear-bottom 96-well microplates (Greiner Bio-One, Kremsmuenster, Austria; Cat#655906). Each well then received 60 µL substrate reaction mix containing: 0.3 mg/mL recombinant human αSyn monomer (Amprion, Inc., San Diego, CA, USA; #S2020), 10 µM thioflavin-T (ThT; MilliporeSigma, Burlington, MA, USA; #T3516), 0.1% sarkosyl (MilliporeSigma, Burlington, MA, USA; Cat#61747), 100 mM PIPES buffer pH 6.5, and 0.4 M NaCl (Sigma Aldrich, St. Louis, MO, USA; Cat#1064040500). Plates were incubated at 42°C in a FLUOstar Omega reader (BMG LABTECH, Ortenberg, Germany) with orbital shaking at 800 rpm for 1 min prior to each of 97 cycles. ThT fluorescence was recorded (excitation 440 ± 10 nm; emission 490 ± 10 nm) after each cycle across 24 h. Data were processed in MARS Omega software v4.00 R2 (BMG LABTECH, Ortenberg, Germany).

For each replicate, the maximum fluorescence intensity (Fmax) served as the basis for classification. As established by previous work [13], replicates with Fmax ≥45,000 RFU were assigned to type 1 α-synuclein seeds; those between 3,000–44,999 RFU to type 2 α-synuclein seeds; and those <3,000 RFU were considered negative. CSF samples with three type 1 replicates were categorized as $S^+$ with type 1 seeds (most often observed in patients with PD/DLB diagnoses). Also in alignment with protocols established earlier [13], samples with two or more type 2 replicates were categorized as $S^+$ with type 2 seeds (most often observed in patients with MSA diagnoses). Samples with two or more negative replicates were categorized as $S^-$. Cases with two type 1 replicates and one type 2 replicate were categorized as $S^+$ of undetermined type, for which repeat testing was recommended. Any other replicate patterns were deemed inconclusive, with repeat testing advised when possible. If repeat testing again yielded an inconclusive or undetermined outcome, that result was retained as the final classification [13]. For consistency, the assay was performed by one experimenter and on one and the same plate reader device throughout the study, and all measurements were performed blinded to clinical diagnosis.

**Statistical analysis**

All analyses were conducted in Python version 3.13.1 (numpy, pandas, statsmodels, scikit-learn) and GraphPad Prism version 10.5.0 (GraphPad Software, San Diego, CA, USA).

Normality and lognormality were assessed by the visual inspection of histograms and Quantile-Quantile plots. Fisher's exact test was used to assess sex differences. Age at visit was compared by Welch's t test; disease duration and global disability scale were compared by log-normal Welch's tests.

NfL values were log-transformed before adjustment for age using ordinary least squares (OLS) regression. Group comparisons of OLS age-adjusted residuals were assessed by the Mann–Whitney U test. Effect sizes were expressed as the rank-biserial correlation (r), calculated according to the formula $r = 1 - 2U/(n_1 n_2)$, using the U statistic provided by GraphPad Prism. For multi-group comparisons, the Kruskal–Wallis test with Dunn's post-hoc correction was applied. Diagnostic performance was assessed by receiver operating characteristic (ROC) analysis. Age-adjusted NfL values were derived using quantile regression ($\tau = 0.5$) and subsequently back-transformed to the original pg/mL scale for reporting. Optimal thresholds were defined by the Youden index within the study cohort and used to dichotomize NfL concentrations as above

($N^+$) or below ($N^-$) the threshold. SynSAA readouts were classified as positive ($S^+$) or negative ($S^-$) for synucleinopathy; inconclusive results, even after a second independent measurement, were excluded. Sensitivity was defined as the percentage of $MSA_{clin}$ cases classified as $N^+$ or $S^+$, and specificity as the percentage of SAOA cases classified as $N^-$ or $S^-$. For the combined biomarker approach (NfL+synSAA), sensitivity was defined as the percentage of $MSA_{clin}$ cases classified as both $S^+$ and $N^+$, and specificity as the percentage of SAOA cases with $S^-$, $N^-$, or both. 95% confidence intervals (CI) of the sensitivity and specificity estimates were computed using the Clopper–Pearson exact method. Area under the curve (AUC) was assessed using z-tests for significance, and bootstrap resampling was used to estimate 95% CI. Positive and negative predictive values (PPV, NPV) were calculated via Bayes' theorem with 95% CI obtained by bootstrapping. As PPV/NPV are prevalence-dependent relative to the given test cohort, clinic-based prevalence estimates of MSA against the background of sporadic adult-onset ataxia were incorporated into the calculations, reflecting the diagnostic context in which these biomarkers are applied. Based on a specialist outpatient cohort [1], MSA accounted for 156 of all 404 patients with sporadic adult-onset ataxia providing a prevalence estimate of 0.4 taken for PPV and NPV computation.

Associations between biomarker readouts and years of disease duration were assessed using Spearman rank correlation, with linear regression models fitted to visualize trends and 95% CI estimated by bootstrapping. Assumptions of monotonic relationship (for Spearman correlation) and linearity, homoscedasticity, and normally distributed residuals (for linear regression) were verified, and outlier influence was evaluated by Cook's distance. For NfL, correlations were performed on age-adjusted residuals obtained from quantile regression. For synSAA, correlations were performed on the median Fmax across three technical replicates per case, as this measure best represented the quantitative readout. Additional analyses restricted to only the MSA-C subgroup were performed for NfL, synSAA, and disease duration correlations to assess consistency and confirmation of the primary findings independent of inclusion of MSA-P cases.

For a stratification into early vs. late disease duration stages, $MSA_{clin}$ subjects were stratified using a 4-year disease-duration threshold (early: <4 years, late: ≥4 years). This threshold was chosen for two reasons: (i) to gauge the diagnostic utility of the biomarkers investigated here, as it has shown–based on natural history and clinicopathological data of MSA–that the diagnostic sensitivity and specificity of the clinical MSA criteria is substantially lower within the first 3 years of disease, representing a critical window in which clinical features may be less fully developed [15]; (ii) to align with disease duration stratification currently deemed most relevant for therapeutic recruitment, reflecting the threshold established in clinical trial precedents and matching the early-stage inclusion criteria of multiple MSA clinical trials (e.g., NCT02315027, NCT02270489).

To estimate the effect of specificity of patient selection on required sample sizes in a hypothetical disease-modifying treatment trial for MSA, we used as exemplary primary outcome the 24-month progression in UMSARS, reported as 21.9 ± 11.9 points/24 months [7]. A fraction (1 – specificity) of patients included in the trial was assumed to be SAOA, modeled as progressing similarly to untreated MSA but not responding to therapy. Assuming equal sizes of treatment and control groups, and equal distribution of SAOA across both arms, the effective treatment effect was derived as

$$TE_{effective} = TE \times \text{specificity},$$

where *TE* is the assumed therapeutic effect in true MSA patients. For simplicity, we assumed equal variance for control and treatment groups. Both specificity values derived against clinical diagnosis (NfL: 0.86; NfL+synSAA: 0.96) were scaled by the literature-reported specificity of the clinically probable MDS MSA diagnostic criteria (0.74) [16], in order to approximate the diagnostic performance expected relative to pathology-confirmed MSA and yield realistic trial sample size estimates. Required sample sizes were then calculated in G*Power (Mann–Whitney U test, two-tailed, α=0.05, power=0.95) using $TE_{effective}$.

#### Data visualization

Schematics were prepared in Biorender™ (BioRender, Toronto, ON, Canada). Plots and statistical visualizations were generated in GraphPad Prism version 10.5.0 (GraphPad Software, San Diego, CA,

USA), and final refinements of figures were performed in Inkscape version 1.4.2 (Inkscape Project; available at www.inkscape.org).

### Supplementary Results

#### Technical validation of the α-synuclein seed amplification assay in an independent cohort

The synSAA discriminates synucleinopathies such as LBD–including PD and DLB–from non-synucleinopathies [13, 17-20], has more recently been applied to MSA [13, 21, 22], and under specific assay conditions (=PIPES-SAA; see Methods), distinguishes type 1 seeds typical of LBD from type 2 seeds characteristic of MSA [13]. To independently validate the PIPES-SAA assay as well as to confirm predefined thresholds for this study in our lab, we first analyzed an independent cohort with the PIPES-SAA protocol leveraging a group of clinically diagnosed LBD ($LBD_{clin}$, n=5), MSA ($MSA_{clin}$, n=10), and non-seeder neurological ward controls (NWC, n=4) that had already been prescreened for synSAA seeding (=technical validation cohort). To determine the gain providing the best clinical group separation, each sample was measured at multiple plate-reader gain settings (range of gain: 700–900). These multiple measurements also allowed to assess consistency of synucleinopathy classification (positive/negative, seed type) across gain settings, based on the maximum fluorescence intensity (Fmax) thresholds (for predefined thresholds, see Methods). For visualization, the median Fmax of three replicates was plotted, as this value reflects the replicate-based classification outcome (**SFig. 1A**). Across all gains, a clear distinction between the three clinical groups was visible: while $LBD_{clin}$ samples showed the highest Fmax values ($LBD_{clin}$/NWC 330.3-392.4; $LBD_{clin}$/$MSA_{clin}$ 3.0-3.2) and controls showed only minimal background signals (58-218 RFU), well below the positivity threshold (3000 RFU). $MSA_{clin}$ samples exhibited distinct intermediate Fmax values ($MSA_{clin}$/NWC 105.6-125.5) across all gains. Based on predefined Fmax thresholds, group distributions showed maximal separation at a gain of 900. At this setting, all 5/5 $LBD_{clin}$ cases (100%) were positive with type 1 seeds, 9/10 $MSA_{clin}$ cases (90%) were positive with type 2 seeds, while one case remained below threshold, and all 4/4 NWC cases (100%) were consistently negative. To further assess separation at the kinetic level, we compared group-level fluorescence traces across gains. Area under the fluorescence curve (AUC) over 24 h showed clearly distinct aggregation kinetics between the three clinical groups: while $LBD_{clin}$ cases showed the highest aggregation kinetics, and NWC remained flat, $MSA_{clin}$ exhibited distinct intermediate aggregation kinetics across all gains **(SFig. 1B)**. Specifically, at a gain of 900, the AUCs were $1.1 \times 10^6$ for $LBD_{clin}$, $2.6 \times 10^5$ for $MSA_{clin}$, and $4.8 \times 10^3$ for NWC **(SFig. 1B)**. Together, these findings provide independent validation that the CSF PIPES-SAA, as also applied here, allows discrimination of synucleinopathy cases from NWC; and in particular detection of the characteristic "intermediate" type 2 seeds in $MSA_{clin}$, distinct from the higher seeding activity in LBD (=type 1 seeds) and the minimal background signals in controls, which can detected based on Fmax and AUC, robustly across gains (**SFig. 1**).

**Supplementary Figures**

**SFig. 1**

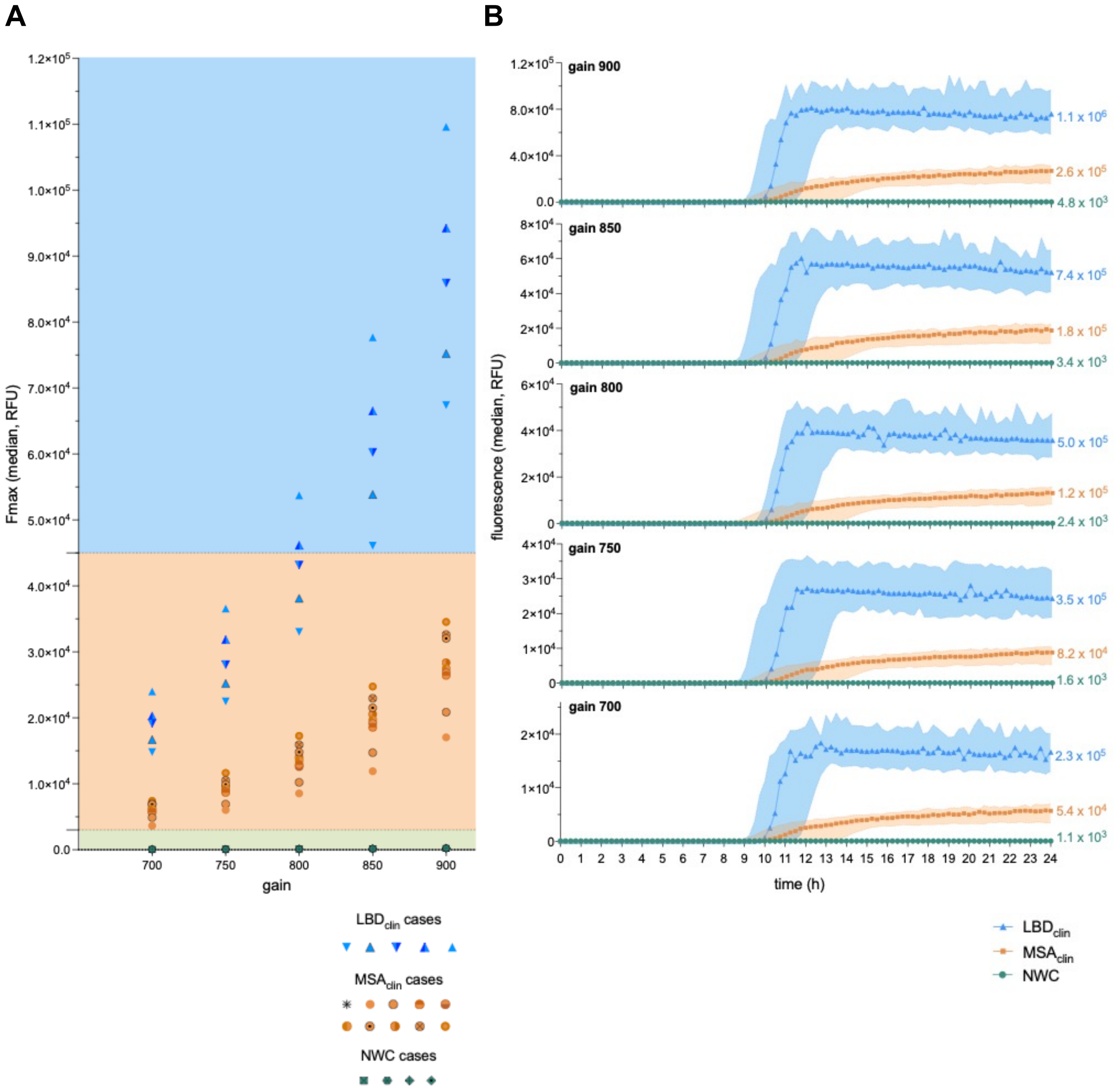


**SFig. 1. PIPES α-synuclein seed amplification assay discriminates $MSA_{clin}$ from $LBD_{clin}$ and neurological ward controls in a validation cohort.** Clinically diagnosed Lewy body disease ($LBD_{clin}$, n = 5), clinically diagnosed multiple system atrophy ($MSA_{clin}$, n = 10), and neurological ward controls (NWC, n = 4) were pre-screened using the PIPES α-synuclein seed amplification assay and selected as a validation

cohort. (**A)** Scatterplot showing maximum fluorescence intensity (Fmax, RFU) at plate-reader gains 700–900, with each datapoint representing one case, shown as the median Fmax of three technical replicate measurements. Shaded regions indicate predefined thresholds: green = negative (<3000 RFU), orange = positive/type 2 (3000 to <45000 RFU), blue = positive/type 1 (≥45,000 RFU). (**B)** Kinetic fluorescence traces (median ± 95% CI) over 24 h at gains 700–900 for each group, with group-level area under the curve (AUC) values shown at the right margin. RFU = relative fluorescence units; CI = confidence interval

**SFig. 2**

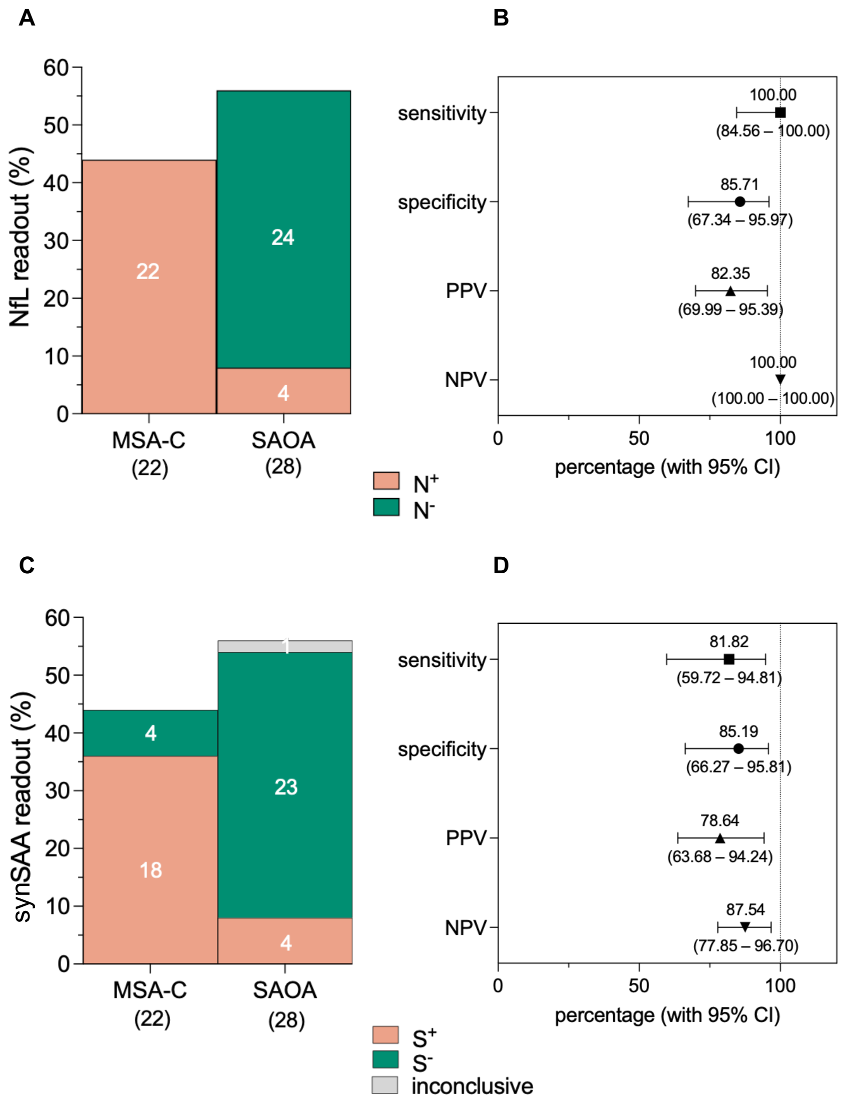


**SFig. 2. CSF NfL and PIPES-SAA discriminate MSA-C from SAOA. (A)** Marimekko plot summarizing the classification of MSA-C (n = 22) and SAOA (n = 28) cases by NfL readout. Readouts were categorized

as NfL-high ($N^+$) and NfL-low ($N^-$), using a threshold (1482.53 pg/mL) determined by the Youden index on age-adjusted residuals in this subgroup comparison. The y-axis indicates the percentage of cases in each category, with case counts shown in white within each stacked bar. **(B)** Forest plot of diagnostic performance metrics for distinguishing MSA-C from SAOA using NfL, showing point estimates and 95% CI. **(C)** Marimekko plot summarizing the classification of MSA-C (n = 22) and SAOA (n = 28) cases by synSAA readout. Readouts were categorized as positive ($S^+$), negative ($S^-$), or inconclusive. The y-axis indicates the percentage of cases in each category, with case counts shown in white within each stacked bar. **(D)** Forest plot of diagnostic performance metrics for distinguishing MSA-C from SAOA using PIPES-SAA, with point estimates and 95% CI. MSA-C = cerebellar-predominant multiple system atrophy (subgroup of $MSA_{clin}$); SAOA = sporadic adult-onset ataxia; NfL = neurofilament light chain; synSAA = α-synuclein seed amplification assay; CI = confidence interval; NPV = negative predictive value; PPV = positive predictive value

**SFig. 3**

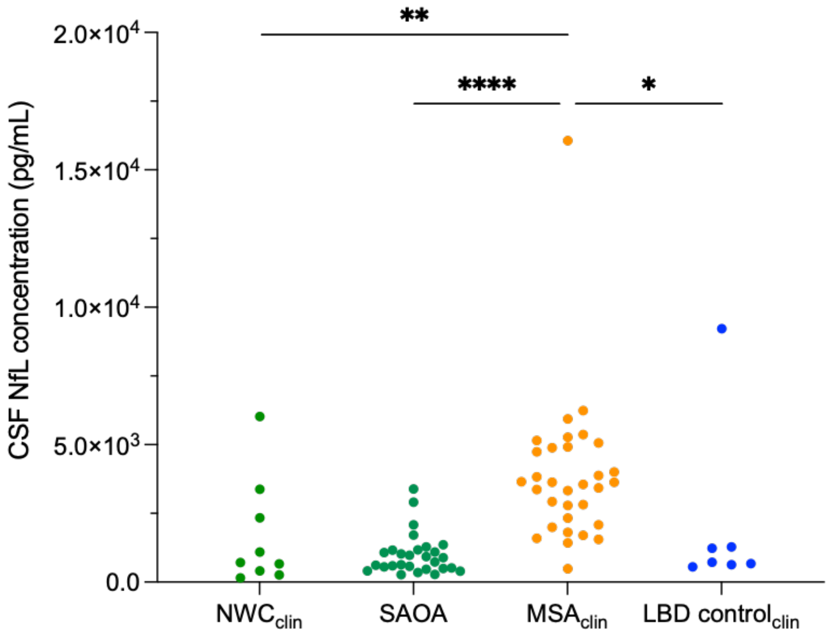


**SFig. 3. CSF NfL concentrations are highest in $MSA_{clin}$ compared with SAOA and clinical reference groups.** Scatterplots of CSF NfL concentrations in the study cohort ($MSA_{clin}$, n = 32; SAOA, n = 28) and clinical reference groups ($NWC_{clin}$, n = 9; LBD $control_{clin}$, n = 4). Each datapoint represents the raw mean NfL concentration of two technical replicate measurements per case. Statistical comparisons were performed on age-adjusted residuals using the Kruskal–Wallis test with Dunn's post hoc correction (*p <

0.05, **$p < 0.01$, ****$p < 0.0001$). $NWC_{clin}$ = neurological ward controls, SAOA = sporadic adult-onset ataxia; $MSA_{clin}$ = clinically diagnosed multiple system atrophy; LBD $control_{clin}$ = clinically diagnosed Lewy body disease; CSF = cerebrospinal fluid; NfL = neurofilament light chain

**SFig. 4**

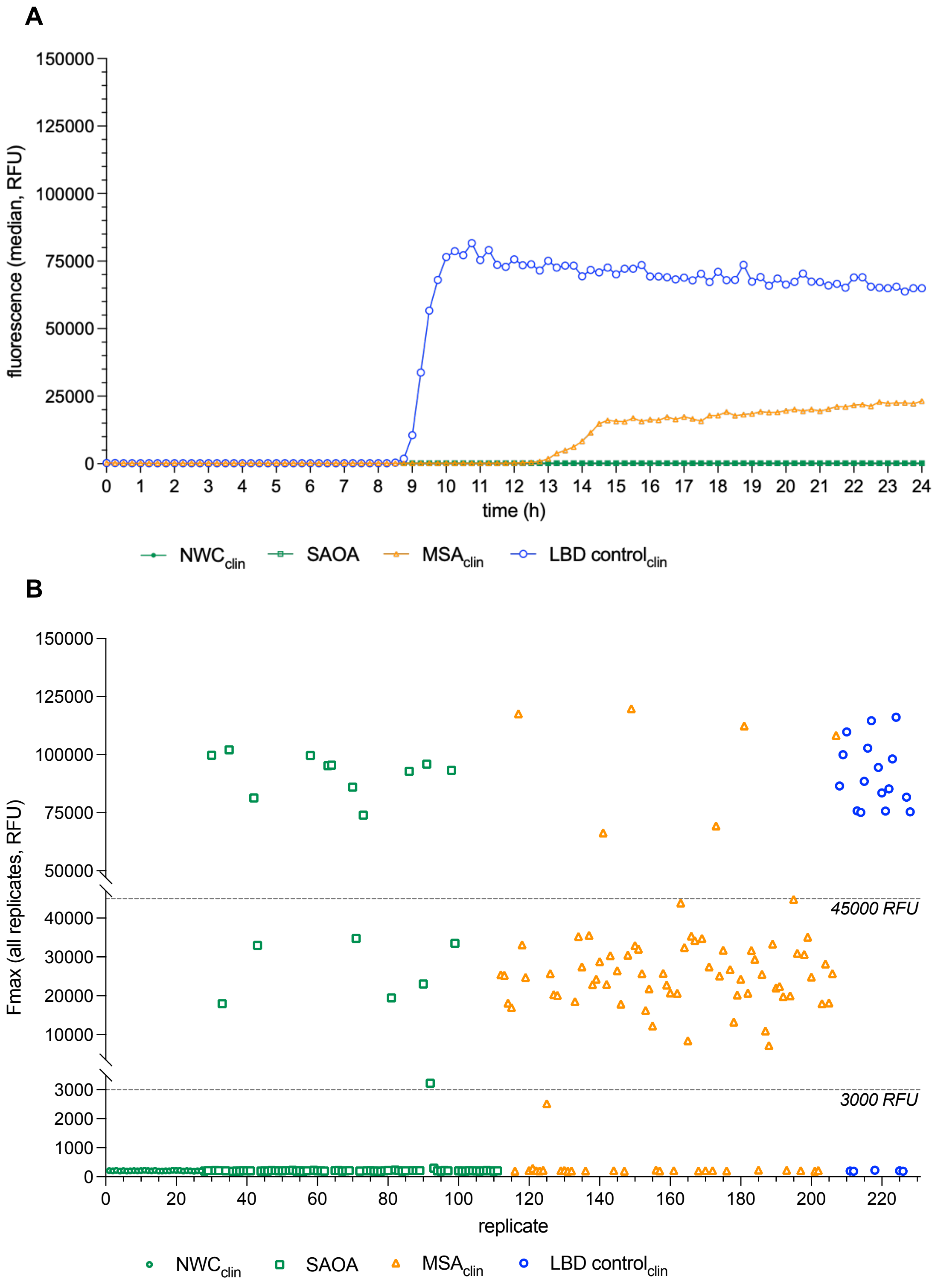


**SFig. 4. PIPES-SAA reveals a distinct seeding signature in MSA$_{clin}$, different from LBD control$_{clin}$ and absent in SAOA and NWC$_{clin}$. (A)** Group-level median fluorescence traces (RFU) over 24 h for LBD control$_{clin}$ (n = 7), MSA$_{clin}$ (n = 32), SAOA (n = 28), and NWC$_{clin}$ (n = 9). Each trace represents the group-

level median, derived from the median of technical replicates within each case. LBD control$_{clin}$ served as a high-seeding clinical reference group, and NWC$_{clin}$ as a negative clinical reference group. (**B**) Scatterplots of all replicate Fmax values (RFU) for each group, showing replicate-level distributions across individual cases. Horizontal dashed lines indicate predefined Fmax thresholds at 3000 and 45000 RFU. Fmax = maximum fluorescence intensity, RFU = relative fluorescence units

**SFig. 5**

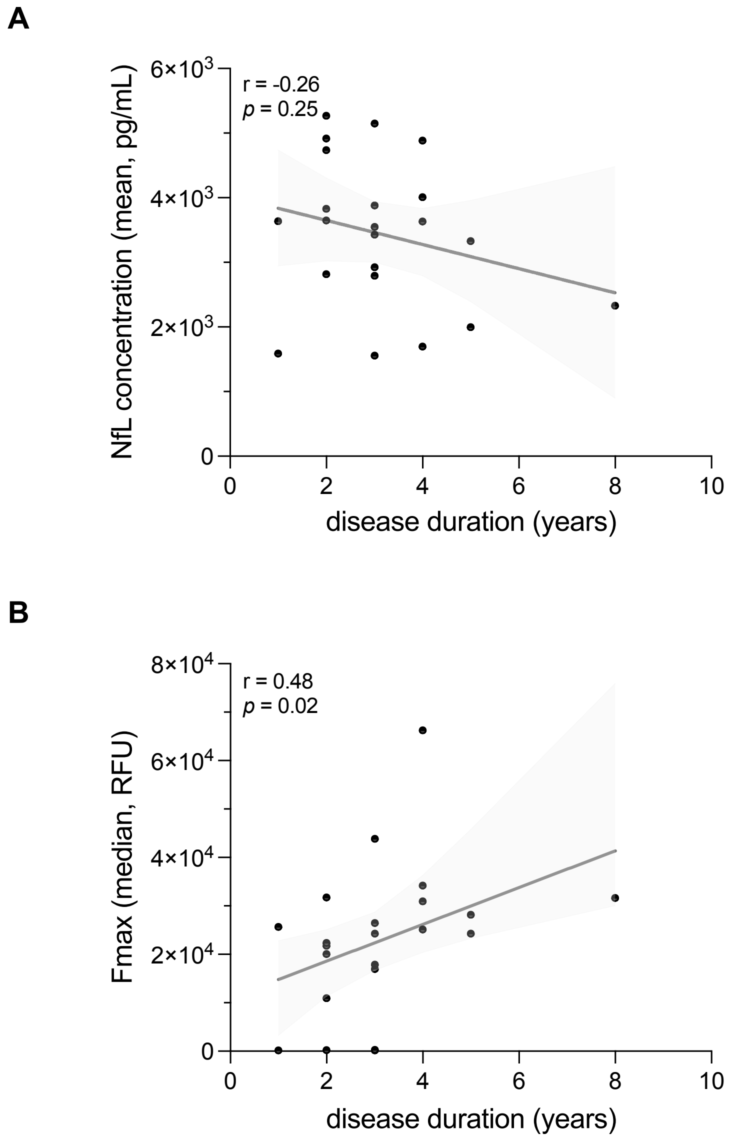


**SFig. 5. Association of CSF NfL and synSAA maximum fluorescence intensity with disease duration in the MSA-C subgroup. (A)** Scatter plot of CSF NfL versus disease duration in the MSA-C subgroup (n

= 22) of MSA$_{clin}$. Each datapoint represents the raw mean NfL concentration of technical duplicates per case. Line (black) shows linear fit and its 95% CI (grey) for visualization. The association was tested using Spearman correlation on age-adjusted residuals (r = – 0.26, p = 0.25). **(B)** Scatter plot of PIPES-SAA maximum fluorescence intensity (Fmax, RFU) versus disease duration in MSA-C cases (n = 22). Each datapoint represents the median Fmax of three technical replicates per case. Line (black) shows linear fit with 95% CI (grey) for visualization; its association is computed by Spearman correlation (r = 0.48, p = 0.024). MSA-C = cerebellar-predominant multiple system atrophy (subgroup of MSA$_{clin}$); SAA = α-synuclein seed amplification assay; NfL = neurofilament light chain; CI = confidence interval; RFU = relative fluorescence units

**SFig. 6**

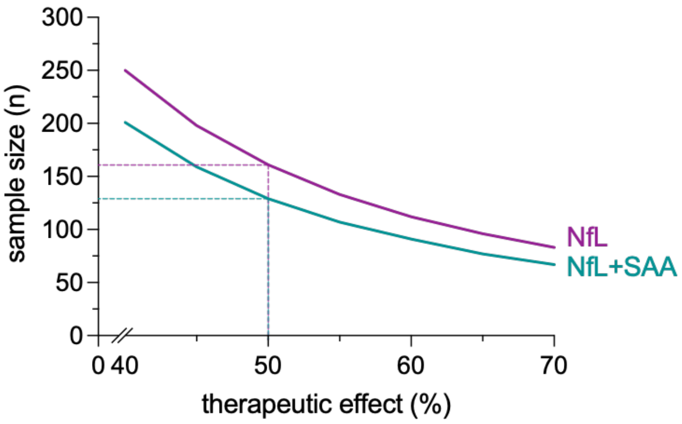


**SFig. 6. Sample size estimates for hypothetical treatment trials based on specificity of NfL versus NfL+synSAA.** Line plots (solid lines) show estimated sample sizes for MSA$_{clin}$ treatment trials based on CSF NfL alone (purple) or combined NfL+synSAA (green), assuming SAOA cases are equally distributed between treatment and control arms. Specificity values (derived against clinical diagnosis) were adjusted based on literature-reported pathology-confirmed diagnostic performance of the clinical MSA criteria reported, to approximate realistic trial sample sizes. The x-axis indicates assumed therapeutic effects (40-70%), and the y-axis the corresponding sample size required. Dashed lines indicate corresponding sample sizes at 50% therapeutic effect. NfL = neurofilament light chain; synSAA = α-synuclein seed amplification assay

### Literature References
